# Racial/Ethnic and Economic Segregation and Survival in the United States

**DOI:** 10.1101/2020.08.21.20179721

**Authors:** I. Mejía-Guevara, M.R. Cullen, S. Tuljapurkar, V.S. Periyakoil, D. Rehkopf

## Abstract

Life expectancy differences across racial/ethnic and economic groups persist in the U.S., but little is known about the combined effects of racial and income segregation in explaining old age survival across neighborhoods. We operationalized neighborhoods using census tracts, with 65,655 of them nested within 3,015 counties, nested within 49 states. We linked census track life table data for ages 45-74 (n=196,965) from the National Vital Statistics System with 5-year population estimates (2011-2015) stratified by race/ethnicity and income from the American Community Survey. We measured racial/ethnic and income segregation using the Index of Concentration of the Extremes at the census tract level, and the indexes of dissimilarity and exposure at the county level. Using three-level random intercept models, we assessed the direct and contextual relationship between survival at ages 45-74 with racial/ethnic and income segregation. Regardless of racial/ethnic stratification, a high concentration of neighborhood poverty was associated with a lower probability of survival relative to affluent neighborhoods (−4.83%; 95% CI −4.86, −4.79), although the relationship was larger in neighborhoods with high concentration of Blacks (−5.61%; −5.67, −5.54). Black-white county-level unevenness also had the largest negative association in those neighborhoods (−0.26%; −0.32, −0.20). Furthermore, Black isolation was negatively associated with a lower survival probability (−0.21%; −0.29, −0.13), but Hispanic isolation was positively associated (0.23%; 0.16, 0.30). Opposite relationships resulted from Black-White (0.06; −0.01, 0.14) and Hispanic-White (−0.13; −0.21, −0.05) interactions. Finally, high exposure to neighborhood poverty/affluence was associated with lower/higher probability of neighborhood survival, but the associations were the strongest for Blacks.

**Significance:** Racial/ethnic and income residential segregation have been associated with negative health outcomes and increased risk of mortality in the U.S. We tested the association between residential segregation at different geographic levels with the survival probability at ages 45-74 across rural/urban neighborhoods. Results indicate that the racial/ethnic and income geographic composition of the population are important factors for explaining this relationship. That is, poverty concentration, minority-white unevenness, and exposure to poverty, had detrimental outcomes for all communities, irrespective of their racial/ethnic composition. However, while minority-isolation/-exposure to whites is detrimental/protective for Black communities, the opposite happened in Hispanic communities. Our research provides important insights in understanding the extent to which residential segregation explains survival disparities across neighborhoods based on their race/ethnicity composition.

## INTRODUCTION

Racial segregation has been regarded for a long time as a fundamental cause of racial disparities in health^1^, rooted in racism, discrimination, and the deliberate use of mechanisms to prevent the interaction of white residents with African Americans or Blacks^2,3^. Income segregation has gained increasing importance in the last forty years as income inequality in the US is growing rapidly. It is contributing to residential stratification where an increasing number of people live in neighborhoods that are either very poor or very affluent, while diminishing middle-class neighborhoods in the process^4^. Racial and income segregation are interrelated as racially segregated neighborhoods are usually correlated with high levels of poverty and deprivation^5,6^, and income segregation has grown more rapidly in recent decades among black and Hispanic families^7^. Here we measured racial/ethnic and income segregation using the Index of Concentration of the Extremes at the census tract level, and the indexes of dissimilarity and exposure at the county level^8,9^.

Residential segregation has been associated with a higher risk of mortality among African Americans, but most studies have been focused on large metropolitan areas, where the great majority of the minority population is concentrated^10–13^. In addition, recent studies have explored trends in the geographic distribution of deaths or survival^14^; by sex and race^15^, socioeconomic status^16,17^, or by cause of death^18^. However, to the best of our knowledge, only a few studies have explored the interaction between racial segregation and economic conditions across neighborhoods in the U.S., mostly based on cross-sectional ecological designs that do not account for small-area conditions^19^.

This study aims to contribute to fill this research gap by examining the combined effects of racial/ethnic and income segregation on old-age survival across neighborhoods in the U.S. We tested the hypothesis that high levels of segregation can aggravate the pervasive effects of neighborhood poverty on the average survival rate. We demonstrated that the racial/ethnic and income geographic composition of the population are important factors for explaining the association between residential segregation and survival at old ages. Specifically, we found that Black/Hispanic isolation was negatively/positivily associated with increased neighborhood survival probability, but the opposite relationships resulted from Black-White and Hispanic-White interactions, respectively. Furthermore, the interaction between racial/ethnic and income segregation, measured by the exposure of minority groups to neighborhood poverty/affluence, was negatively/positively correlated with neighborhood survival. Finally, we further examined the interaction between income concentration at the tract level and racial/ethnic unevenness at the county level, and found a larger gap in the probability of survival between poor and rich neighborhoods with high concentration of Blacks in counties whith high racial unevenness.

We followed a multilevel approach to account for variation and clustering across different geographic levels, as well as for a proper assessment of contextual effects of segregation, while accounting for county-level risk factors traditionally associated with high mortality risk across racial/ethnic groups, as well as rural/urban differences and small-area geographic variation.

## METHODS

### Data and Study Participants

Data for this study are from 65,655 census tracts, nested within 3,015 counties, and 49 States (including the District of Columbia, but excluding Maine, Wisconsin, and U.S. Territories). We operationalized census tracts as neighborhoods, which are relatively stable statistical subdivisions of a county with an average of 4,000 inhabitants^20^. It is the main unit of analysis for this study that includes life table estimates at this geographic level for people aged 45 and older. Data are reported in 10 age groups: 45-54, 55-64, 65-74, 75-84, and 85+, but we excluded the age groups 75-84, and 85+, to avoid any problems in mortality rate estimates in sparse older groups, as well as to allow focus on premature mortality^21,22^, which leaded to 196,965 total observations in our analytic sample.

### Outcome

The outcome for this study is the probability of survival between age groups *a* and *a+n* (S_a+n_), conditional on reaching ages 45 to 65 (e.g., the probability of surviving from age 45 and 54 is represented by S_45–54_). We computed survival rates from life table values of L_a_ (the number of person-years lived between ages *a* and *a+n*) using the expression: 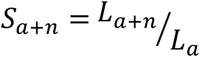 (e.g., the probability of surviving to age 54 conditional on reaching age 45 is given by 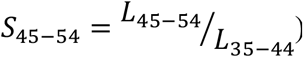^23^.

We computed survival rates for 65,655 census tracts across the U.S. using abridged life period tables from the U.S. Small-area Life Expectancy Project (USALEEP), that were constructed by combining standard demographic techniques and statistical modelling at the census tract level^24^. They relied on death counts from the National Vital Statistics System (NVSS) and the National Center for Health Statistics (NCHS) occurring between 2010 and 2015, and population counts from the 2010 decennial census and the 2011-2015 American Community Survey (ACS) 5-year estimates. Life tables include adjustments that account for errors in population estimates, leading to very good reliability when compared to national estimates^25^. Finally, it is worth pointing out that these life table estimates are not disaggregated by race/ethnicity/sex so results may reflect differences in population composition of the neighborhood, but not racial/ethnic/sex differences in survival.

### Exposures: Racial and Income Segregation

The main exposures for this study are racial/ethnic and income segregation, which were measured at two different levels: census tract and county. With tract level exposures, we were able to measure direct associations between probability of survival and census tract residential segregation. Meanwhile, county-level segregation exposures allowed the testing of cross- or contextual-level associations.

Neighborhood Income and Racial Stratification

Income concentration is measured at the tract level using the Index of Concentration of the Extremes (ICE), introduced by Massey^9^ to quantify how persons in a specified area are concentrated into the poor vs. affluent groups of the income distribution. We classified families in each neighborhood as poor (P) based on the 2015 Federal Poverty Line (FPL) of $24,250 (we used the threshold of < $25K in 2015 dollars) for a family of 4 persons^26^, while affluent (A) groups in the neighborhood as those who earned $75K or more, which is the threshold of the 5_th_ quintile of the national household income distribution at the census tract level. ICE was computed using the formula:

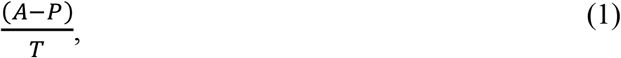

where T represents the total population with reported income in the same neighborhood, and A, P, and T were obtained from the ACS 2011-2015 5-year estimates^20^. We denoted the resulting index as ICE_i_, where *i* is a referent to income. ICE_i_ is then a continuous measure that ranges from −1 to 1, which connote 100% concentration of poor and affluent, respectively, but for statistical analysis, we stratified ICE in tertiles (classification of neighborhoods in three groups, each comprising nearly one third of the ICE ordered distribution).

Similarly, we use ICE to measure racial/ethnic concentration^27^, by replacing P and A in formula 1 by grouping persons representing deprived and privileged groups, respectively. However, instead of considering non-Hispanic Blacks (hereinafter Blacks) and non-Hispanic Whites (hereinafter Whites) as representing deprived and privileged groups as in previous studies^27^, we included Hispanics in the computation of ICE for a better assessment of the stratification of census tracts that considers the three largest racial/ethnic groups in the U.S.

We performed this calculation by applying the ICE formula twice. That is, ICE was computed for Whites and Other race/ethnicity (step 1) and then stratified in tertiles (step 2). We excluded the third tertile that comprised the majority of white populations from the previous grouping and calculated ICE again in the remaining tracts with Hispanics (representing the privileged) and Blacks (representing again the most deprived) as reference groups (step 3). The resulting ICE was split in two groups of equal size that was appended to the previously excluded tertile from the second iteration to generate a three-group index that we denoted as ICE_r_ (the subscript *r* refers to race/ethnicity) (step 4) (SI Appendix, **Figure S1**).

County Racial and Income Residential Segregation

At the county level, racial/ethnic residential segregation was estimated using the Index of Dissimilarity (_x_D_w_) between minority and white groups, which is a widely used index of racial evenness that measures the percentage of minority and white populations that would need to be redistributed across census tracts to achieve an even residential distribution^1,2,8^, and it was computed using the formula:

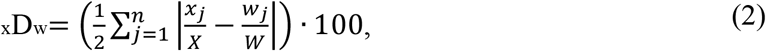

where *x_j_* in the number of minority members in tract *j*, and *w_j_* the number of Whites in the same tract, *n* is the number of tracts in the county, and X and W are the total populations of each group in the county^8^. This index varies from 0 to 100, indicating complete integration or segregation, respectively, and values of this index above 60 are viewed as “high” whereas those below 30 are considered as “low”^2^. To isolate the effect of distinct minority groups in our statistical analysis, we defined _x_D_w_ separately for Blacks (_b_D_w_), Hispanics (_h_D_w_), and non-Whites (hereinafter Other,_o_D_w_).

A separate measure of racial segregation was conducted using the index of exposure, that considered two distinct dimensions: minority isolation and minority interaction with white residents in the same neighborhood. To measure this kind of segregation, we used the within-tract interaction index (_x_P_y_), which reflects the likelihood that a minority person *x* shares a unit area with a majority person *y*. We computed _x_P_y_ using the formula:

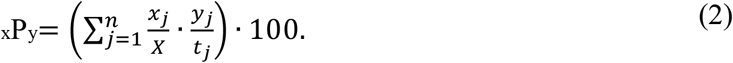

where *x_j_* and X are defined as before, *y_j_* represent the number of majority group in the tract and *t_j_* is the total population in tract *j*. _x_P_y_ also varies from 0 to 100 representing the probability that a minority person shares a unit area with a majority person^8^. Specifically, to estimate minority isolation (e.g., Black isolation) *y_j_* in formula 2 is replaced by *x_j_* (and it is denoted as _x_P_x_), *y_j_* represents the number of Whites in the neighborhood in the estimation of minority-White interaction (_x_P_w_).

Similarly, we used equation 2 to assess county-level income segregation by measuring the potential exposure of minority groups to neighborhood poverty/affluence^28^. In this case, we replace *y_j_* in equation 2 by the number of poor(p)/affluent(a) residents in the neighborhood, respectively. For ease of interpretation, values of _x_P_p_ (minority-poor interaction) and _x_P_a_ (minority-affluent interaction) below 20 or above 40 are interpreted as low/extreme concentration levels of poverty/affluence, respectively.

In our statistical analysis, both measures _x_D_w_ and _x_P_y_ were standardized to have mean zero and standard deviation of one in each state to account for differences in the distribution of both indexes across states and to facilitate interpretation of point estimates.

### Sensitivity Analysis: Adjusting for county-level risk factors as suggestive of mechanisms

In this study, we incorporated the prevalence of three well-known predictors of mortality at the county level: adult smoking, physical inactivity, and excessive drinking. These factors have been identified in the past as the three leading contributors of mortality in the U.S., accounting for nearly 40% of total deaths^29^, but we only included them as suggestive of mechanisms for sensitivity analysis. These prevalence estimates were obtained from the 2017 County Health Rankings & Roadmaps project, which collects a comprehensive set of health and behavioral estimates at the county level across time in the US^30^, and estimated using survey data from the Behavioral Risk Factor Surveillance System (BRFSS). The prevalence of Adult smoking represents the percentage of the adult population (aged 18 and older) in a county that reported currently smoking every day or most days and have smoked at least 1,000 cigarettes in their lifetime. Physical inactivity refers to the percentage of people aged 20 and older reporting no leisure-time physical activity in the past month obtained from three years of survey responses from BRFSS. Excessive drinking measures the percentage of adult population in the county reporting binge or heavy drinking in the past 30 years and was obtained from 1 year of survey data from BRFSS. All these factors were also standardized to have mean zero and standard deviation of one in each state in our statistical models.

Although previous studies have indicated that BRFSS prevalence rates were comparable to other national surveys, limitations persist due to question differences among those surveys used for comparison. This is particularly relevant to the predictors of alcohol consumption and physical inactivity, as it could bias results if other surveys had been used to estimate the prevalence for this study^31^.

### Statistical Analysis

We fitted three-level random intercept models to assess the relationship of survival between ages 45-74 with the racial/ethnic and income residential segregation. We used two different model specifications to test the separate effect of racial and income segregation at the county level and its interaction with neighborhood racial and income concentration. The first model specification (hereinafter Model A) was as follows:

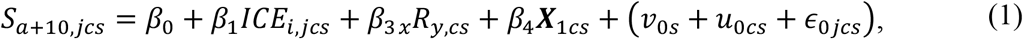

where *S*_*a*+10,*jcs*_ is the probability of survival from age *a* to *a*+10 conditional on reaching age *a* (45, 55, and 65) in neighborhood *j*, within county c, within state *s*. Similarly, *ICE_i,jcs_* is the index of income concentration at the respective levels. *_x_R_y,cs_* represents the index of racial/ethnic segregation at the county level, where separate models where used depending on the type of racial/ethnic segregation. That is, we used the index of dissimilarity, *_x_R_y,cs_* = *_x_D_w,cs_*, to assess minority-white unevenness between minority *x* and Whites (*w*) in county *c*, within state *s*; and the index of exposure, *_x_R_y,cs_* = *_x_P_y,cs_*, to assess minority isolation (*_x_P_x_*), or minority-white interaction (*_x_P_w_*). ***X***_1*cs*_ is a vector with tract-level covariates that includes age specified by 10-year groups, and rural/urban neighbor defined as tracts with a population density of less/more than 1000 persons per square mile^28^. The three last terms in the right-hand side of equation 1 are respectively the random intercepts errors at the state (*v*_0*s*_), county (*u*_0*cs*_), and neighborhood (*∊*_0*jcs*_) levels, which are assumed to be normally distributed with mean 0 and standard deviations, *σ_v_*, *σ_u_*, and *σ_∊_*. We conducted national level as well as stratified models by racial stratification using tertiles of the ICE_r_ index for a better assessment of the racial geographic variation across the country.

The second model specification (hereinafter Model B) was used to examine the effects of income segregation at the county level using the interaction index *_x_P_y_* as a proxy for minority interaction with poor/affluent individuals within the same neighborhood. The model specification is as follows:

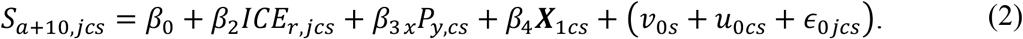

In equation 2, *ICE_r,jcs_* is the index of racial concentration in neighborhood *j*, within county *c*, in state s; *_x_P_y,cs_* stands for the percentage of *y* (Poor/Affluent) individuals residing in the same neighborhood of the minority group *x*, and ***X***_1*cs*_ is defined above. As in Model A, we conducted stratified analysis by levels of neighborhood racial concentration using tertile groups of ICE_r_.

Finally, we conducted sensitivity analysis by adding the 3 county-level factors’ prevalence defined above (adult smoking, physical inactivity, and excessive drinking) into the specifications defined by equations 1 and 2. The objective of this sensitivity analysis is testing the robustness between the potential association between residential segregation and survival, rather than looking at the mechanisms mediating those associations, which is beyond the scope of this research.

## RESULTS

### Population Concentration in Neighborhoods by Race/Ethnicity and Income

Racial/Ethnic and Income Concentration

**Figure 1** illustrates the racial and income composition of ICE exposures at the tract level. Panel A shows the distribution of ICE_r_ by race/ethnicity, where groups 1-3 concentrate 78%, 79%, and 47% of the Black, Hispanic, and White population at the national level (hereinafter denoted as Black/Hispanic/White Concentrated groups), respectively. The within-group composition shows that Group 1 and 2 comprised around 50% of the minority population, where the majority is represented by Blacks (30%) in Group 1 and Hispanics (38%) in Group 2. Meanwhile, the great majority are Whites (92%) is Group 3. Panel B illustrates the respective national and within group distribution of ICE_i_, with Group 1 concentrating 49% of the poorest households while Group 3 is comprised with 57% of the affluent. Within Group 1, 39% of households were poor and 16% affluent, in contrast to Group 3 where 11% were poor and 56% were affluent.

**Figure 1.**
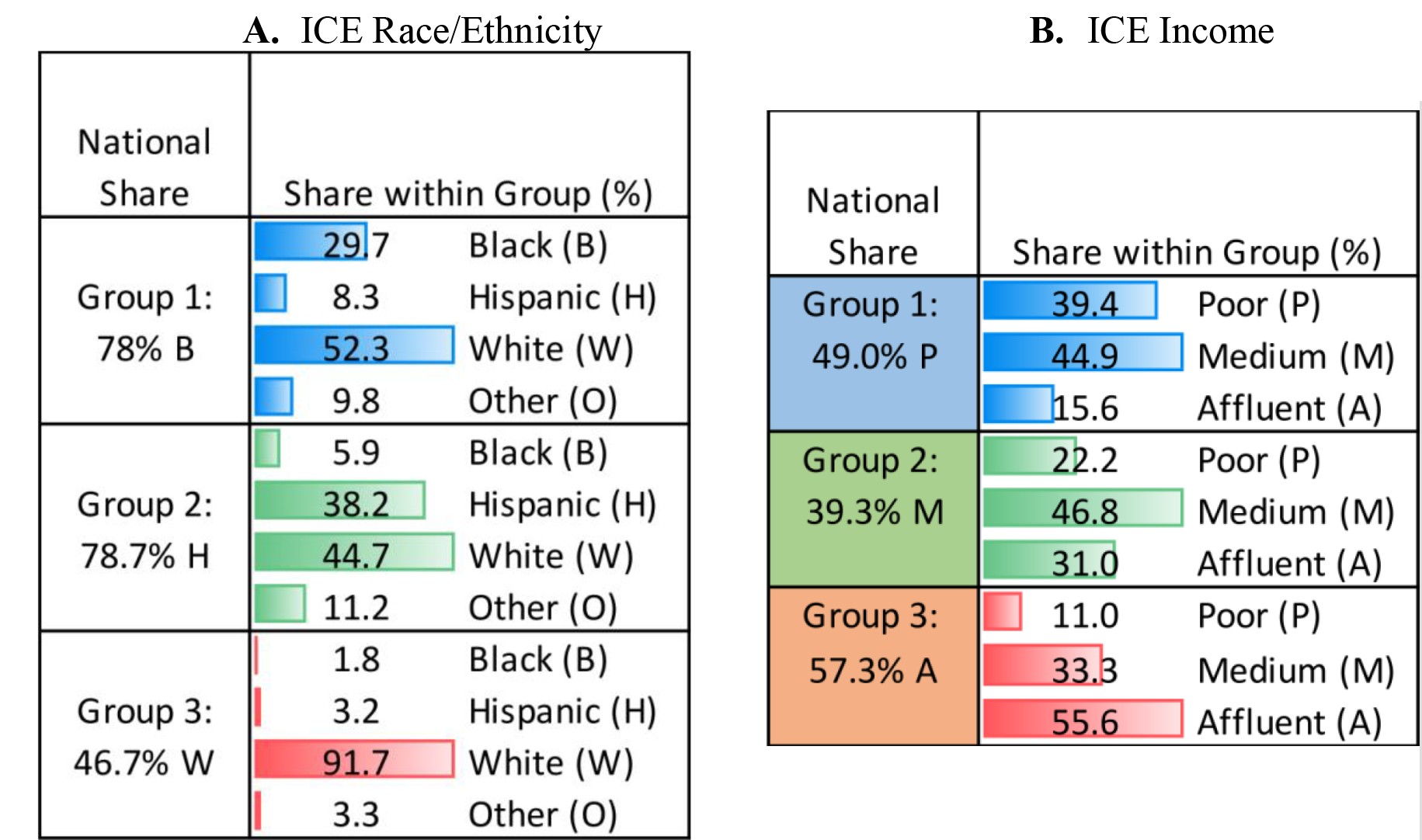
Tract-level exposures: Index of Concentration of the Extremes (ICE) and population share based on race/ethnicity (A) and income (B).

**Note:** In panel B, Poor refers to households (HH) with less than $25K, Medium includes HH income between $25-$74K, and Affluent by HH with income ≧ $75K.

Racial Stratification and geographic Distribution

**Figure 2** illustrates the geographic distribution of each group in ICE_r_ by census tract, where clear patterns emerged reflecting the racial/ethnic composition of the three predominant groups, Blacks, Hispanics, and Whites. Panel A shows that neighborhoods with a high concentration of Blacks are mostly situated in the south-western states, the east coast, and in large metropolitan areas in the east north central and California. Panel B indicates that Hispanics are concentrated in the West coast, South (Texas and Florida), and with important presence in large metropolitan areas in the Northeast (NY/NJ), and East North Central (Chicago) regions. Finally, the neighborhoods with the highest concentration of Whites are located mainly in the north and center, from the Pacific to New England, and in some states in the south (Arizona, Texas, Florida).

**Figure 2.**
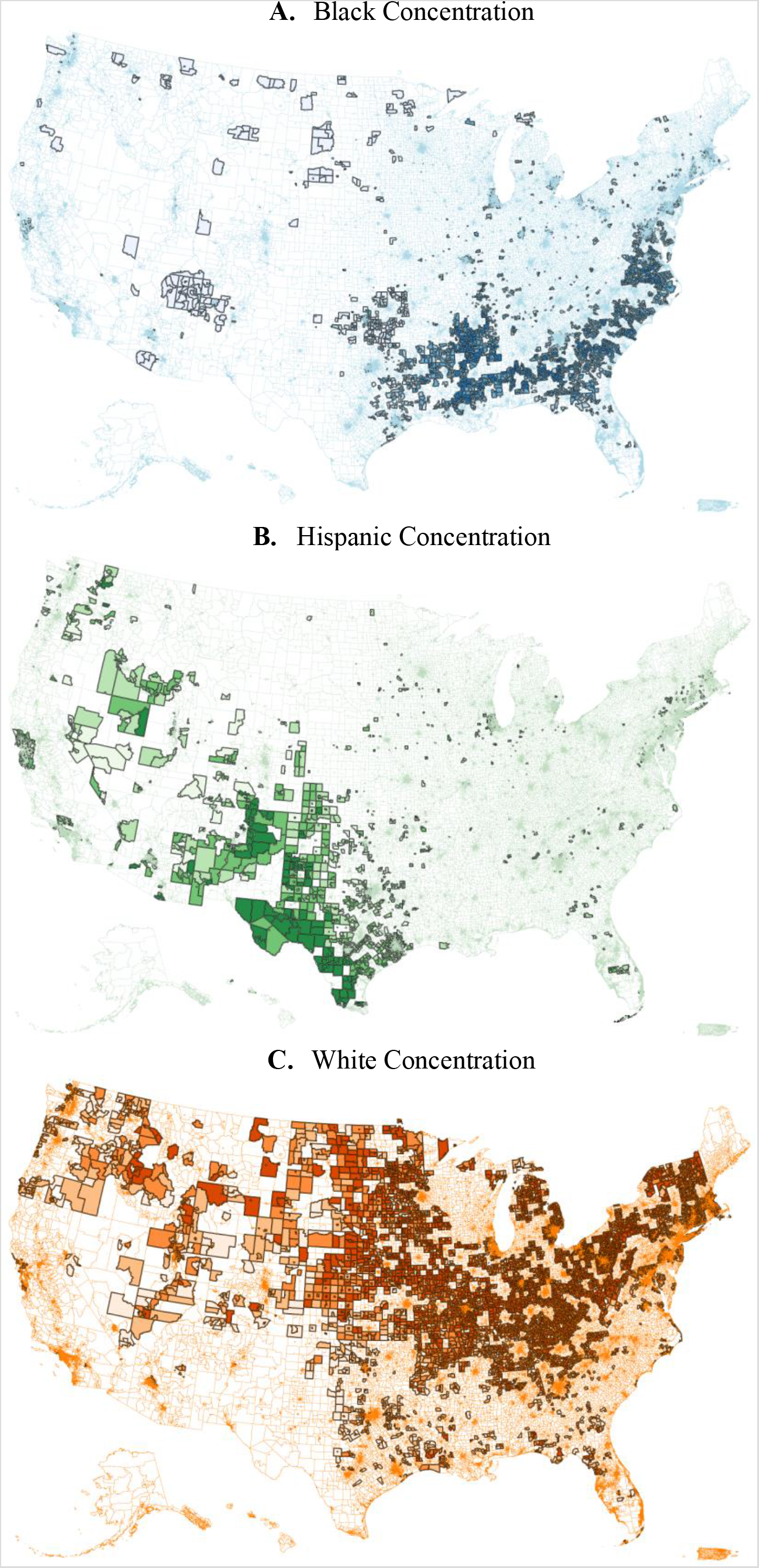
Geographic distribution of ICEr in tracts stratified by race/ethnic concentration of Blacks (A), Hispanics (H), and Whites (C) from ICEr in the US, 2010–2015.

**Source** Authors’ with data from 2011–2015 ACS 5-year estimates.

We used this geographic stratification in our subsequent analysis to differentiate associations of residential segregation with survival probabilities in groups of neighborhoods where a specific minority group is more represented.

### County-level Racial/Ethnic and Income Segregation

We show in **Table 1** the distribution of county-level segregation reflecting evenness and potential exposure to neighborhood poverty/affluence, stratified by ICE_r_ or race/ethnic concentration.

**Table 1.** Distribution of county-level residential segregation by racial unevenness, minority isolation and interaction with Whites, and exposure to neighborhood poverty/affluence, in stratified models by race/ethnic concentration.

|  | Neighborhood Racial/Ethnic Stratification |  |  |
| --- | --- | --- | --- |
|  | Black<br>Concentration | Hispanic<br>Concentration | White<br>Concentration |
| **Dissimilarity (xDw), median (IQR)** | **Black-White** | **Hispanic-**<br>**White** | **Other-White** |
| National | 52.6 (20) | 46.4 (16.2) | 33.0 (18.6) |
| Poverty | 54 (21.7) | 45.9 (17.8) | 29.3 (16.3) |
| Medium | 51.1 (19.2) | 44.8 (17.6) | 31.9 (18.2) |
| Affluence | 52.2 (17.3) | 48.4 (12.3) | 36.6 (17.4) |
| **Minority Isolation and interaction with**<br>**Whites, median (IQR)** | **Black** | **Hispanic** | **Other** |
| Minority Isolation (xPx) | 37.2 (32.7) | 43.3 (28.9) | 23.3 (30.6) |
| Interaction with Whites (xPw) | 41.4 (30.4) | 35.1 (27.4) | 76.7 (30.6) |
| **Racial-Income Interaction, median (IQR)** | **Black** | **Hispanic** | **White** |
| Exposure to Poverty (xPa) | 12.0 (5.6) | 8.2 (2.7) | 8.8 (4.0) |
| Exposure to Affluence (xPa) | 8.8 (4.7) | 9.5 (3.3) | 13.1 (5.6) |

**Note:** IQR stands for the inter-quartile range of the distribution in the respective stratified region and group.

Compared to Whites, we observe that Blacks living in highly concentrated Black neighborhoods are less evenly distributed [median _b_D_w_ = 52.6, interquartile range (IQR) = 20.0] than Hispanics [46.4 (16.2)] and Other minorities (33.0 [18.6]) in the respective Hispanic and Whites concentrated regions. We observed a gradient across income-stratified neighborhoods in Black regions, with larger segregation in those with a higher concentration of poverty [54.0 (21.7)] than affluence [52.2 (17.3)]. This pattern is less consistent, however, for other racial/ethnic groups (**Table 1**).

**Table 1** also shows that Blacks in highly concentrated Black neighborhoods are more likely to be exposed to neighborhood poverty [median _b_P_p_ = 12.0, IQR = 5.6] than Hispanics [8.2 (2.7)] and Whites [8.8 (4.0)], and also less exposed to neighborhood affluence [8.8 (4.7)] than Hispanics [9.5 (3.3)] and Whites [13.1 (5.6)].

### Association of racial/income residential segregation and old-age survival

Model A. Neighborhood concentration of income and racial segregation

Results for model A indicate that differences in income concentration had significant negative effects on the neighborhood probability of survival in average. Compared to neighborhoods with high concentration of affluence, the most deprived neighborhoods had a lower probability of survival of 4.83% (95% CI: −4.86, −4.79) in average (**Table 2**). In addition, a one standard deviation increase in racial segregation at the county level was also negatively associated with a lower probability of survival of −0.19% in average (−0.22, −0.15). In racially stratified tracts, the associations of neighborhood concentration varied significantly across regions. In Black concentrated neighborhoods, the difference in survival probability in poor neighborhoods—relative to affluent ones—was significantly higher (−5.61%; −5.67, −5.54) than the corresponding poor neighborhoods with high concentration of Hispanics (−3.75; −3.80, −3.69) or Whites (−4.13; −4.19, −4.07). Black and Hispanic residential unevenness relative to Whites were also significantly associated with reduced neighborhood survival probability of −0.26% (−0.32, −0.20) and –0.11% (−0.18, −0.03) in their respective regions (**Table 2**).

**Table 2.** Model A. Multilevel linear association between probability of survival between ages 45-74 and neighborhood income concentration and county racial segregation in the U.S. and by racially-/ethnic-stratified regions

|  |  | Racial/Ethnic Tract Stratification |  |  |
| --- | --- | --- | --- | --- |
|  | National | Black<br>Concentration | Hispanic<br>Concentration | White<br>Concentration |
| Tract: Income Concentration (ICEi) |  |  |  |  |
| Group 1 (Poor) | -4.83<br>[-4.86,-4.79] | -5.61<br>[-5.67,-5.54] | -3.75<br>[-3.80,-3.69] | -4.13<br>[-4.19,-4.07] |
| Group 2 | -2.06<br>[-2.09,-2.03] | -2.31<br>[-2.38,-2.25] | -1.74<br>[-1.79,-1.69] | -1.92<br>[-1.96,-1.87] |
| Group 3 (Affluent: referent) | 1.00 | 1.00 | 1.00 | 1.00 |
| County: Racial/Ethnic Segregation (xDw, z-Score) |  |  |  |  |
| Other/White (oDw) | -0.19<br>[-0.22,-0.15] |  |  | -0.04<br>[-0.08,-0.00] |
| Black-White (bDw) |  | -0.26<br>[-0.32,-0.20] |  |  |
| Hispanic-White (hDw) |  |  | -0.11<br>[-0.18,-0.03] |  |
| Covariates |  |  |  |  |
| Urban Neighborhood (Rural: referent) | -0.48<br>[-0.51,-0.44] | -0.33<br>[-0.40,-0.26] | -0.37<br>[-0.43,-0.31] | -0.36<br>[-0.41,-0.32] |
| Age Group (45-54: referent) | 1.00 | 1.00 | 1.00 | 1.00 |
| 55-64 | -3.34<br>[-3.37,-3.31] | -3.90<br>[-3.96,-3.85] | -3.29<br>[-3.34,-3.25] | -2.83<br>[-2.87,-2.79] |
| 65-74 | -9.75<br>[-9.78,-9.73] | -10.87<br>[-10.92,-10.81] | -9.58<br>[-9.63,-9.54] | -8.82<br>[-8.86,-8.78] |
| N observations | 196470 | 65316 | 65334 | 65820 |
| N census tracts | 39294 | 13063.2 | 13066.8 | 13164 |

95% confidence intervals in brackets.

We tested the interaction between income concentration at the tract level with county-level racial unevenness, and found that in concentrated Black neighborhoods, the probability of surviving at age 65-74 was significantly lower in poor neighborhoods when the counties exhibit high racial segregation (panel A of **Figure 3**); however, in Hispanic concentrated neighborhoods survival probabilities were lower in poor neighborhoods compared to affluent ones, but it declined in high income neighborhoods within counties with high levels of segregation (panel B of **Figure 3**). The latter is partly explained by rural/urban differences in survival, which is significantly lower in urban as compared to other settings (**Table 2**). Finally, the probability gap between poor and affluent neighborhoods did not increase significantly in segregated places compared to less segregated ones (panel C of **Figure 3**). Minority isolation and interaction with Whites

**Figure 3.**
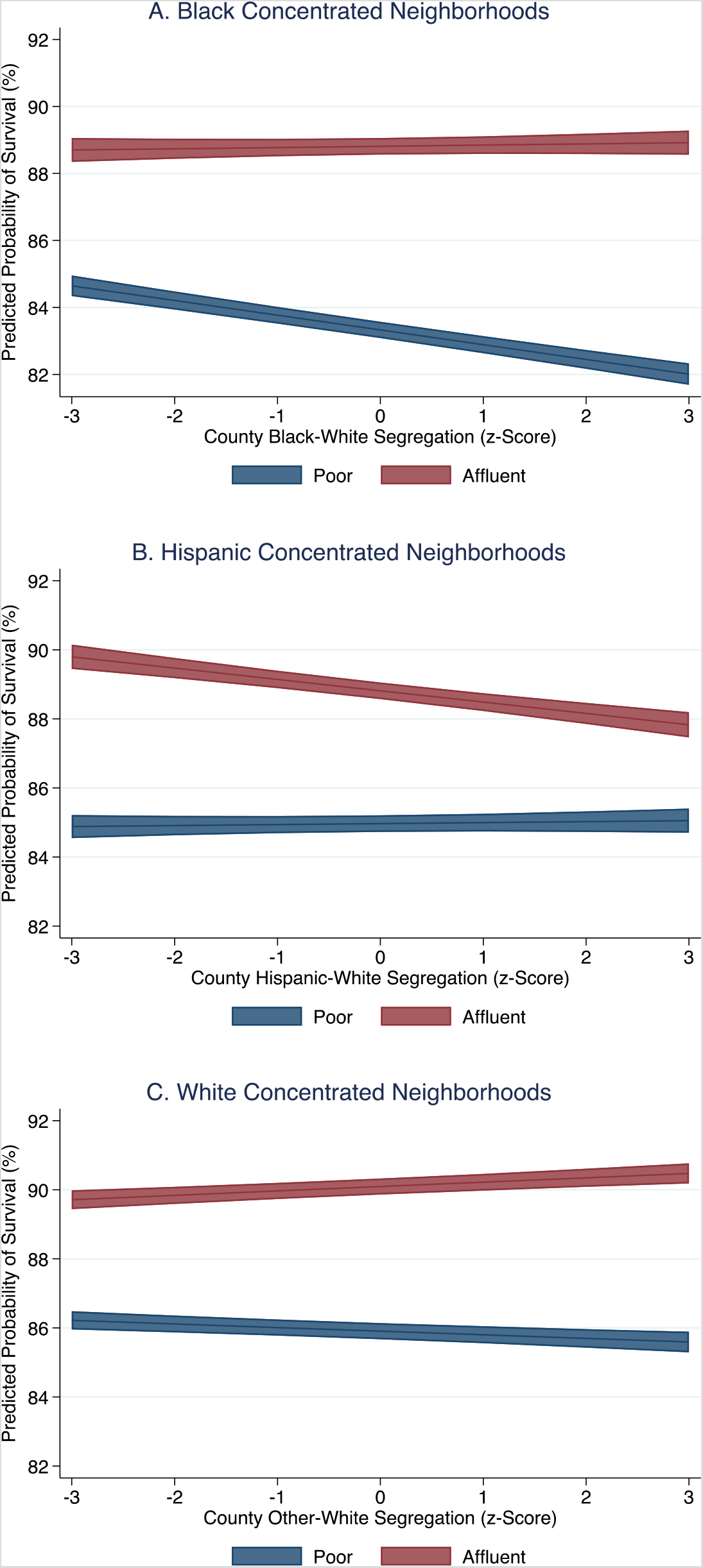
Neighborhood by county predictive probability of survival at age 65-74 in neighborhoods with high concentration of Blacks (panel A) and Hispanics (panel B).

**Note:** Predictive values were obtained by including the interaction term ICEi*IDr in equation 1 (Model A). Models were stratified by neighborhoods with high concentration of Blacks (panel A), Hispanics (panel B), or Whites (panel C). We controlled for age groups (45-54, 55-64, and 65-74) and rural/urban tract.

Our results of minority isolation are reported in **Figure 4**. We found that while for Blacks exposure to other black minorities or isolation was negatively associated with a lower probability of survival (−0.17%; −0.25, −0.10), isolation among Hispanics seems to be protective and leaded to an increase in the neighborhood survival probability (0.24%; 0.16, 0.31) (panel A of **Figure 4**; SI Appendix, **Table S1**). In contrast, the reverse was observed with a higher exposure of minorities to white residents, since the average survival probability of neighborhoods increased with a larger Black-White interaction (0.06%; −0.01, 0.14), but decreased with a larger Hispanic-White interaction (−0.13%; −0.21, −0.05) (panel A of **Figure 4;** SI Appendix, **Table S1**).

**Figure 4.**
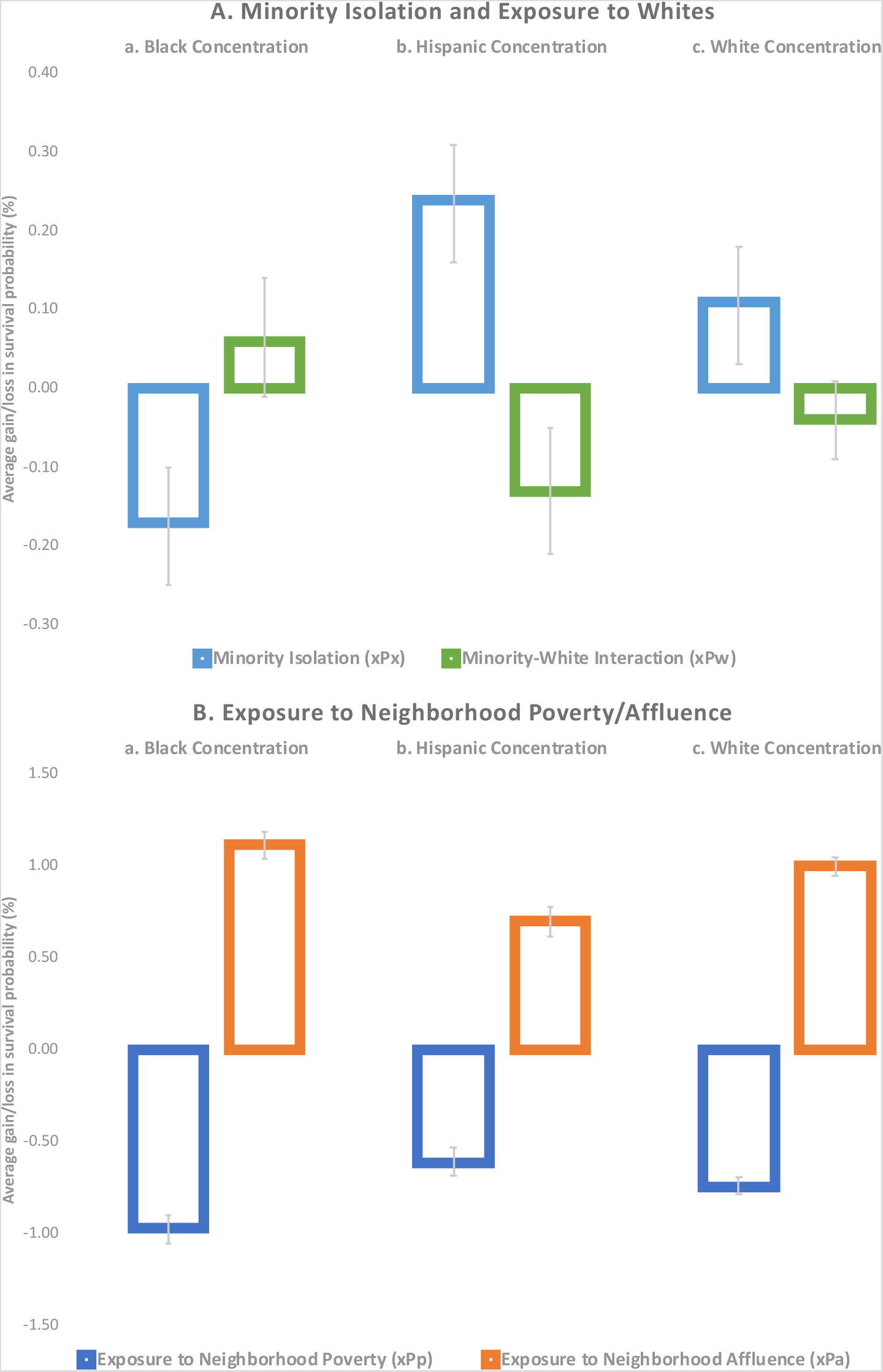
Contextual associations between tract probability of survival between ages 45-74 and county residential segregation, measured by minority isolation and interaction with Whites (panel A), and economic segregation measured by the interaction of race/ethnic residents with neighborhood poverty/affluence (panel B), in stratified models by race/ethnic concentration.

**Note:** Estimates were obtained using the model specification of equation 2, where we controlled for age groups (45–54, 55–64, and 65-74) and rural/urban tract. Point estimates for all exposures and risk factors are in **Table S1** in the Supplementary Information.

Model B. Neighborhood racial concentration and exposure to poverty and affluence

Panel B of **Figure 4** summarizes the main contextual associations between neighborhood survival probability and county level exposure to poverty/affluence in stratified models by racial/ethnic concentration. Irrespective of the racial/ethnic concentration, a higher exposure to neighborhood poverty had a negative association in the neighborhood probability of survival but increased with a higher exposure to neighborhood affluence. However, the effect size was higher for Blacks in Black concentrated neighborhoods than the exposure of Hispanics and Whites in their respective strata. A one standard deviation increase in the exposure to neighborhood poverty among Blacks, reduced the average probability of survival of the neighborhood by 0.97% (95% CI: −1.05, −0.90)—a significantly larger association than for Hispanic (−0.61%; −0.68, −0.53) and White (−0.74; −0.78, −0.69) concentrated neighborhoods, respectively—, while it increased with a one standard deviation increment in the exposure to neighborhood affluence by 1.12% (1.04, 1.19)—also significantly larger than in the respective Hispanic (0.70%; 0.62, 0.78) and White (1.00%; 0.95, 1.05) concentrated neighborhoods—(panel B of **Figure 4;** SI Appendix, **Table S1**).

Sensitivity Analysis to inclusion of risk factors

After the inclusion of the county prevalence of smoking, physical inactivity, and alcohol drinking into the specification of Model A, the association between the probability of survival and minority-white unevenness was not affected (SI Appendix, columns xDw of **Table S2**). For instance, in neighborhoods with high concentration of Blacks, black-white unevenness had the same association with the probability of survival [−0.27% (−0.33, −0.21) in **Table S2** compared to −0.26% (−0.32, −0.20) in **Table 2**). In addition, all of the county risk factors had significant negative contextual associations with the neighborhood probability of survival. For instance, in neighborhoods with high concentration of Hispanics, one standard deviation increase in the prevalence of smoking, reduced physical activity, and excessive drinking, reduced the average probability of survival of the neighborhood by −0.27% (−0.36, − 0.18), −0.30% (−0.39, −0.20), and −0.34% (−0.43, −0.26) in average, respectively (SI Appendix, column hDw of **Table S2**). Similarly, the association between minority-isolation and interaction with whites also remained consistent, with a slightly change in the effect size (panel A of **Figure 4**; SI Appendix, panel A of **Figure S2**). In the case of model B, the association between minority exposure to poverty/affluence was attenuated by around 40% after adjusting for county risk factors in all models, although the associations remained statistically significant (panel B of **Figure 4**; SI Appendix, panel B of **Figure S2**).

## DISCUSSION

This study has 4 main findings. First, we show that it is important to account for the geographic distribution of the population by racial/ethnic groups for a proper assessment of the contextual effects of residential racial segregation in the U.S. A clear clustering of racial/ethnic groups emerged that is rooted by a historical process of residential segregation and migration patterns across small geographies throughout the country^7,28,32,33^. Previous studies have mainly focused on the analysis of residential segregation within metropolitan areas, given the important concentration, large number, and diversity of minority populations in these areas. Our study is more comprehensive because it looks at small-area segregation across the whole country as it relates to the survival of older populations and captures important variation across urban and rural settings. It also departs from pure ecological studies by accounting for contextual associations of residential segregation on the probability of survival within neighborhoods while accounting for variation at the tract, county, and state levels^34–36^.

Second, this study showed that segregation that is characterized by the unevenness of racial/ethnic groups had significant contextual associations with the probability of survival in neighborhoods with high concentration of Blacks or Hispanics. Our results indicate that income is an important predictor of neighborhood differences in survival, which echoes extensive studies showing relationships between income and inequality with life expectancy^14,15,36–38^, but the stronger effects across racial groups are better appreciated when a proper stratification of racial/ethnic geographic units is conducted.

Third, we further assessed the association of minority isolation and interaction with Whites. We found a significant negative association of Black isolation with neighborhood survival, which echoes previous findings regarding racial segregation as a fundamental cause of racial disparities in health^1^, and associated with a higher risk of mortality among African Americans^10–13^. In contrast, we found that Hispanic isolation had protective effects as reflected by a larger probability of survival in regions with high concentrations of Hispanics. This result may be a reflection of the social support hypothesis proposed in the literature to explain the so-called “Hispanic Paradox”—where survival among foreign-born Hispanics experienced lower mortality and better health in several outcomes relative to non-Hispanic whites despite their lower socioeconomic status and access to healthcare^40^. According to this hypothesis, Hispanics possess stronger family ties that help them build a sense of community and potentially contributing with a better health^41^, although our results should be interpreted with caution because our life table data only reflect neighborhood differences as they are not disaggregated by racial/ethnic group. Finally, our index of interaction showed that a larger Black-White interaction—e.g., less segregation—was positively associated with a higher probability of survival in the neighborhood, but the Hispanic-White iteration was negatively associated. Although the mechanisms behind this finding are unclear and beyond the scope of this research, this may be attributed to differences in racial-/ethnic-income composition, a larger exposure of discrimination and racism, or operating through socioeconomic restrictions imposed to members of minority groups (e.g., restricting access to education and employment opportunities)^2,42,43^.

Fourth, this study was particularly designed to examine cross-level interactions of income and racial/ethnic segregation. We used the index of exposure to examined the interaction between distinct racial and economic groups for the assessment of income residential segregation^28^. In particular, we looked at the association between the probability of survival and the exposure of race/ethnic groups to neighborhood poverty and affluence. Our results indicate that exposure to poverty/affluence was negatively/positively correlated with neighborhood survival, but the effect size was significantly higher in Black communities compared to those with high concentration of Hispanics or other minorities, particularly with regard to the exposure to poverty. This particular finding stresses the importance of economic exposure to improve the survival in Black communities in particular, given the recent evidence demonstrating that Blacks are more/less likely exposed to poverty/affluence^28^.

Future research using individual mortality data and predictors by race/ethnicity should shed further light on this important finding. Finally, we further examined the interaction between income concentration at the tract level with racial/ethnic unevenness at the county level. We confirmed significant differences in survival probabilities between poor and affluent neighborhoods, where the gap surged with increasing residential segregation at the county level in neighborhoods with high concentration of Blacks. Although the gap in the predictive probability of survival between poor and affluent neighborhoods remained in regions with high concentration of Hispanics, the effect of residential segregation reduced the survival gap rather than the opposite as shown in Black concentrated neighborhoods, which may be attributed to rural/urban differences in survival as well as to higher life expectancy of Hispanics compared to Whites. As we stated before further research will be needed to examine the mechanisms behind this finding.

Our sensitivity analysis revealed that the associations of residential segregation and neighborhood survival were robust to variations in the county-level prevalence of smoking, physical inactivity, and excessive drinking, aggregates of important behavioral predictors of mortality at the individual level^29,44–46^. Although the association of exposure to poverty/affluence was attenuated in the presence of these factors, they remained significant in all the stratified models. While there are many more risk factors for mortality that could be considered, it shows that our primary findings are not completely explained by a small number of the strongest behavioral risk factors for mortality.^44–48^ It is worth noting that these are not confounders in the traditional sense since they do not cause segregation themselves, and they are more likely to be mediators, and further research may be required to look at the mechanisms.

### Limitations

This study has three main limitations. First, life table data were not available by race/ethnicity, hence our conclusions should be interpreted as contextual level effects potentially affecting the overall probability of survival in neighborhoods rather than any specific race/ethnic group associations. Second, extensive studies have shown that gender differences in life expectancy, particularly across racial/ethnic groups, are important^44,49^, but our data does not account for such differences because the lack of information disaggregated by sex. Thirdly, our control for the three risk factors for mortality is only suggestive of mechanisms, since this is an incomplete measure of the pathways through which these factors may mediate the relationship between residential segregation and survival. As such, we do not formally test for statistical mediation.

## Conclusions

This study showed that residential segregation, as represented by racial/ethnic unevenness and exposure, are important predictors of neighborhood disparities in the survival across regions with high concentration of minority groups in the U.S. In particular, residential segregation remains negatively pervasive for Black populations, not only because it remains the most segregated racial group, but also because residential segregation of Blacks is particularly detrimental for their survival at older ages when accompanied with high exposure to poverty. Our results, however, show that minorities, specially Blacks, can overcome these pervasive effects when they are exposed to high neighborhood affluence, so infrastructure or other focalized projects that increase economic opportunities to minority communities may contribute in reducing the racial gaps in survivorship at old age in the U.S.

## Data Availability

N/A

## Acknowledgement

Research reported in this publication was supported by the National Institute On Aging of the National Institutes of Health under Award Number P30AG059307. The content is solely the responsibility of the authors and does not necessarily represent the official views of the National Institutes of Health. The corresponding author had full access to all the data and had final responsibility to submit the paper for publication. Data for this project were accessed using the Stanford Center for Population Health Sciences Data Core. The PHS Data Core is supported by a National Institutes of Health National Center for Advancing Translational Science Clinical and Translational Science Award (UL1 TR001085) and from Internal Stanford funding.

## Supplemental Information

**Table S1.**
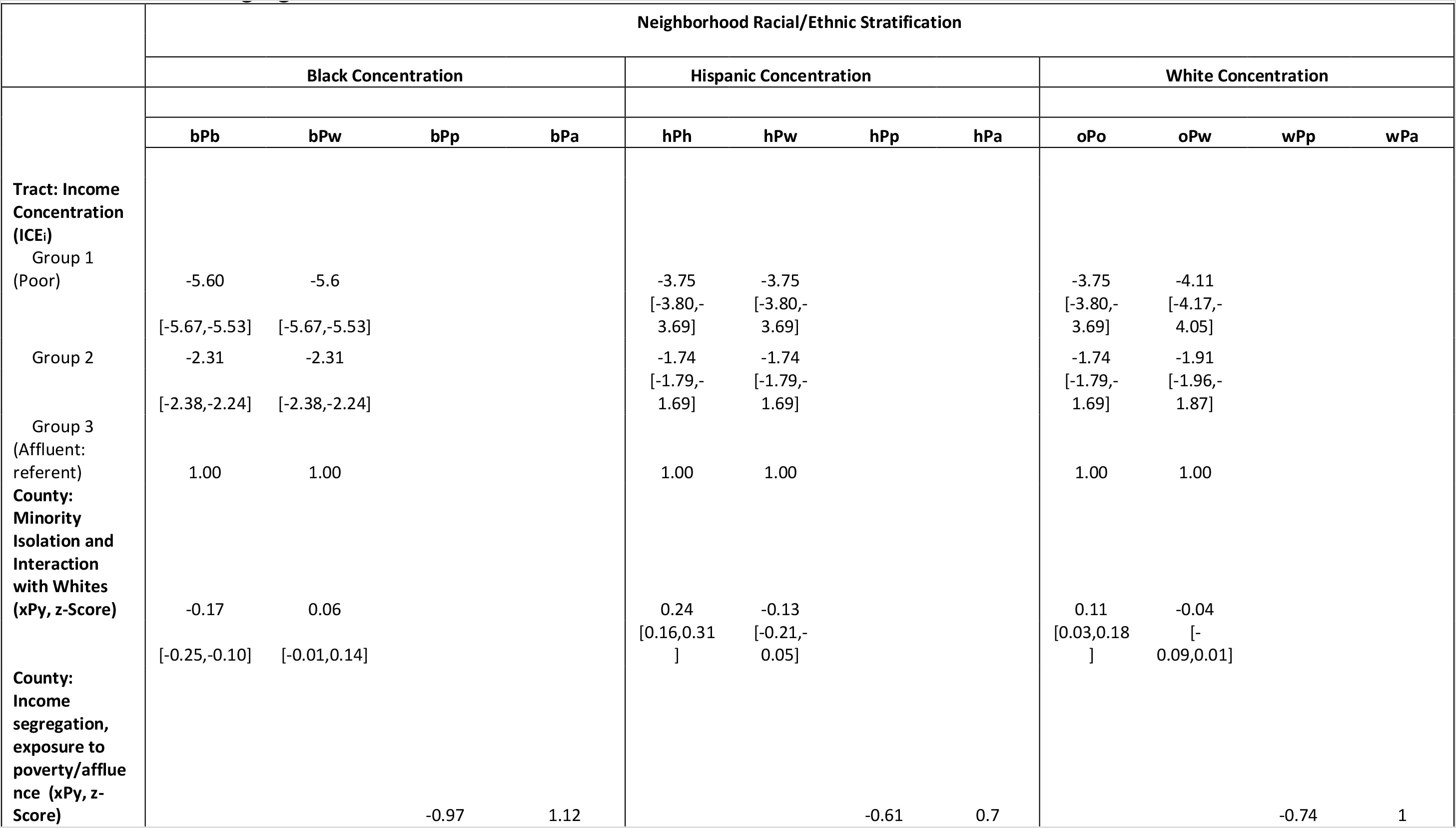
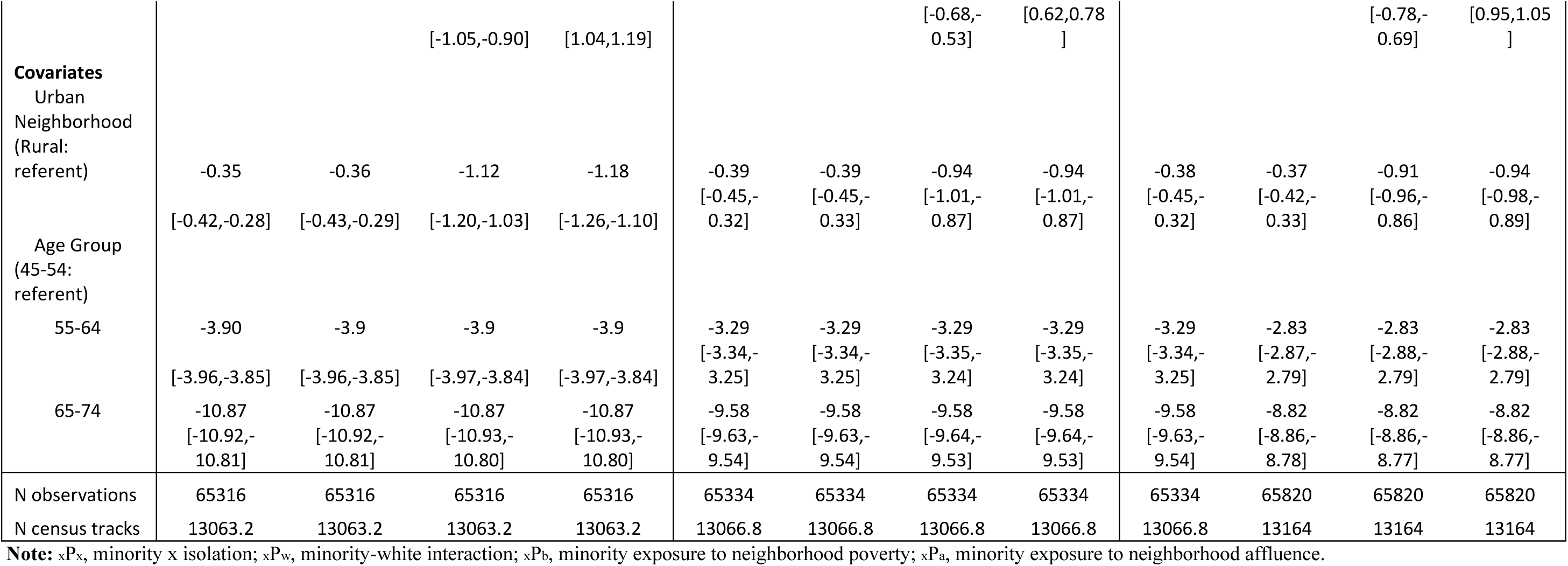
Model B. Multilevel linear association between probability of survival between ages 45-74 with neighborhood racial and income segregation in stratified models based on racial/ethnic concentration

**Table S2.**
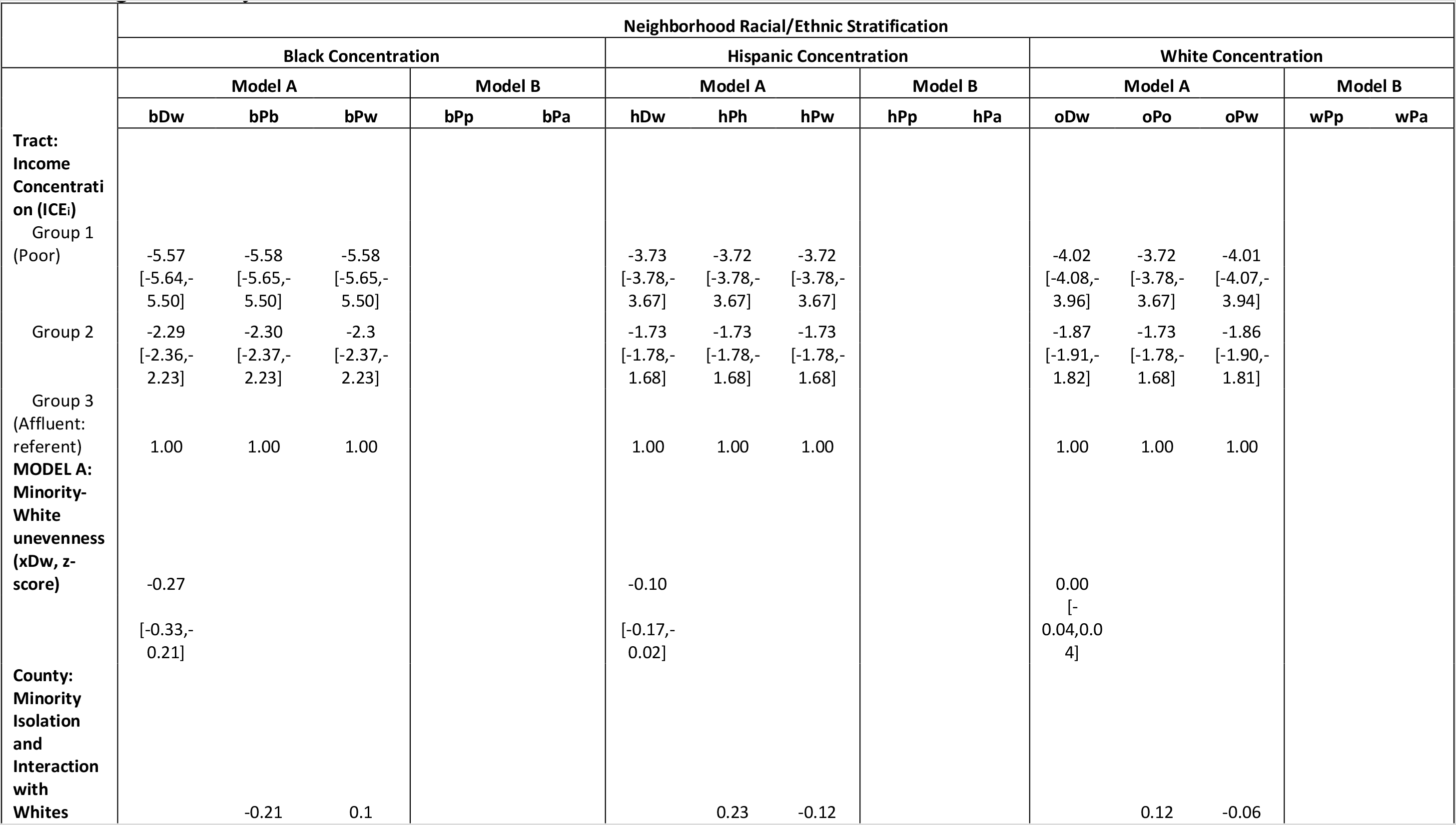
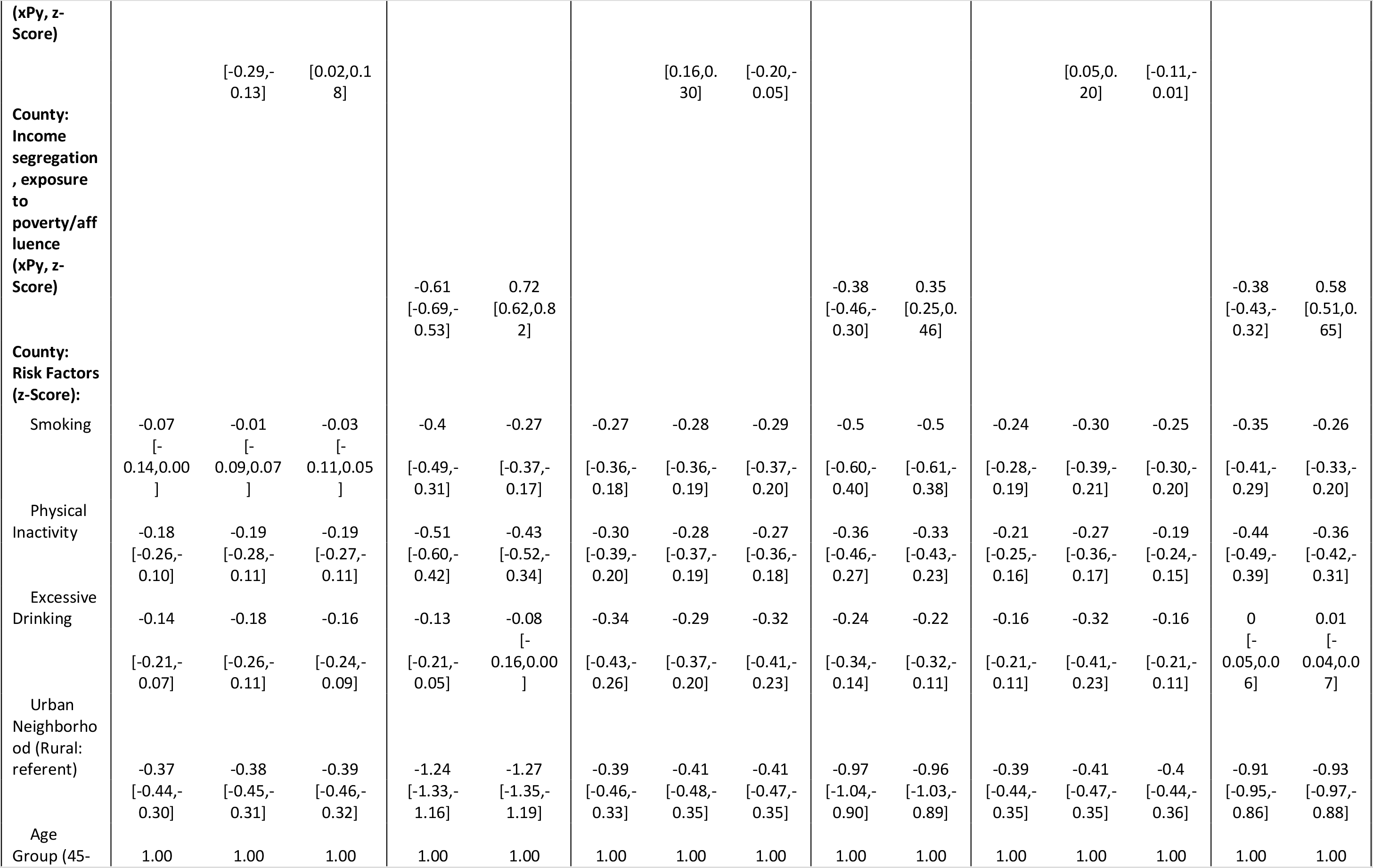
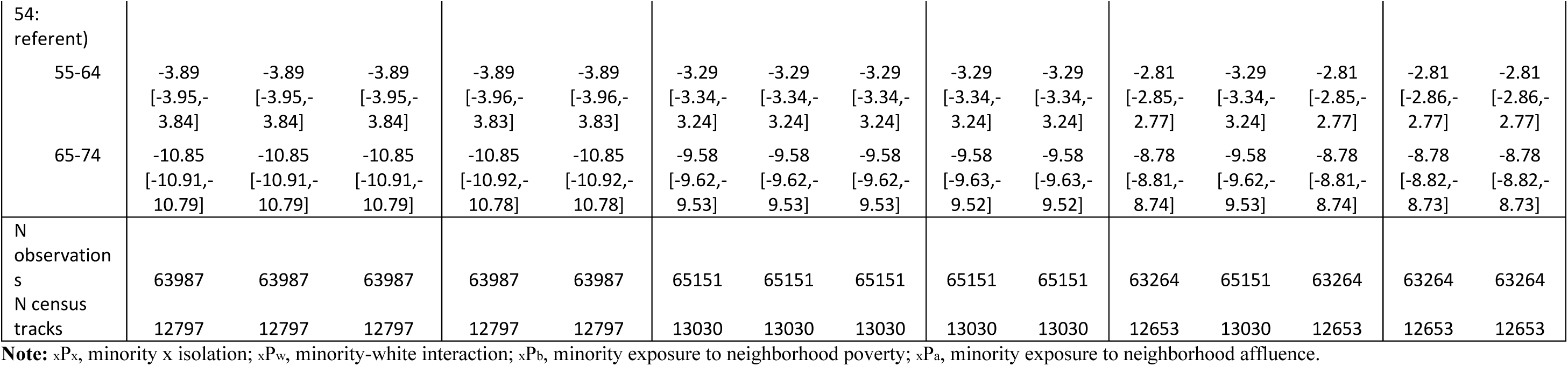
Sensitivity Analysis. Multilevel linear association between probability of survival conditional between ages 45–74 with neighborhood racial and income segregation in stratified models based on racial/ethnic concentration, and after accounting for county level risk factors

**Figure S1.**
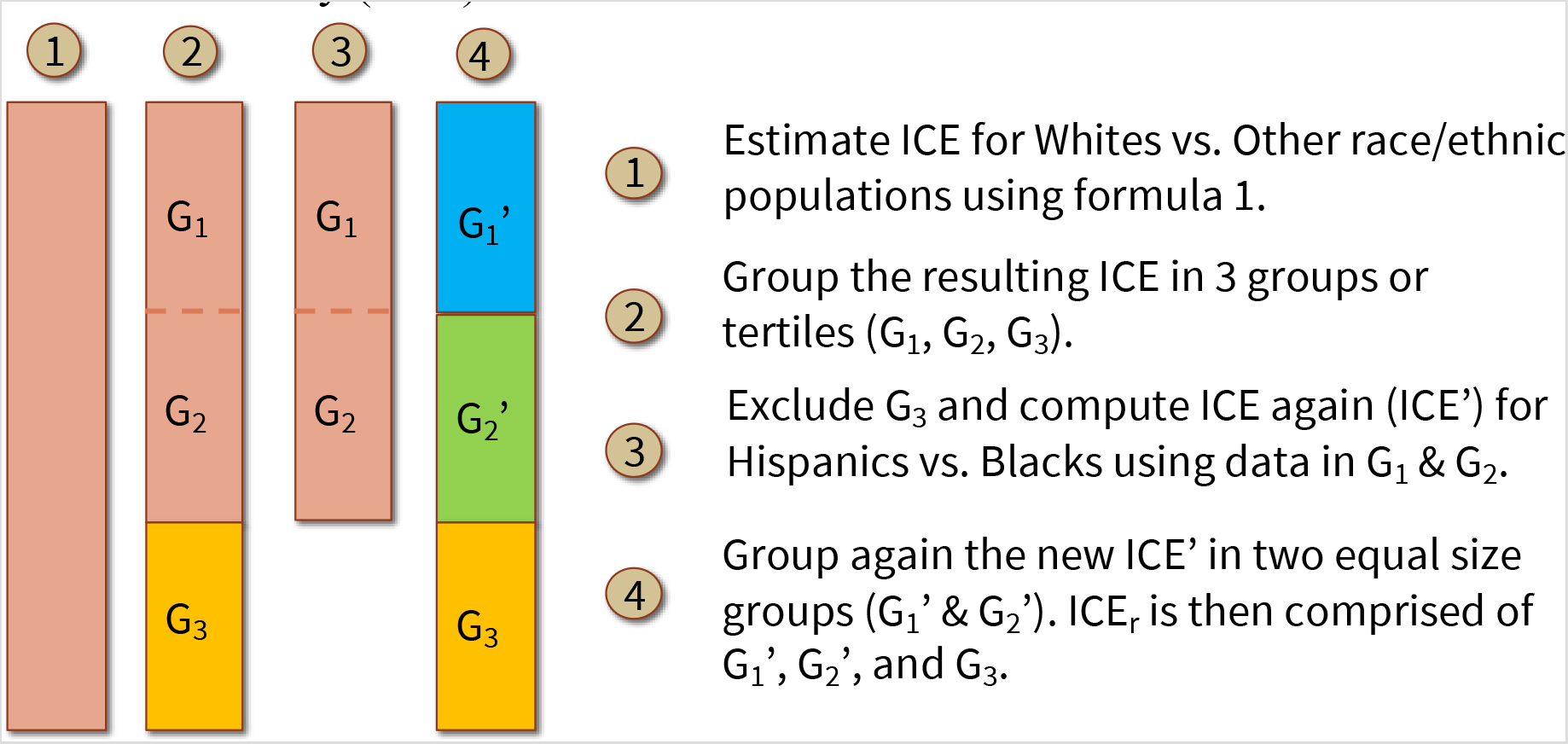
Procedure to estimate the Index of Concentration of the Extremes for race/ethnicity (ICE_r_)

**Figure S2.**
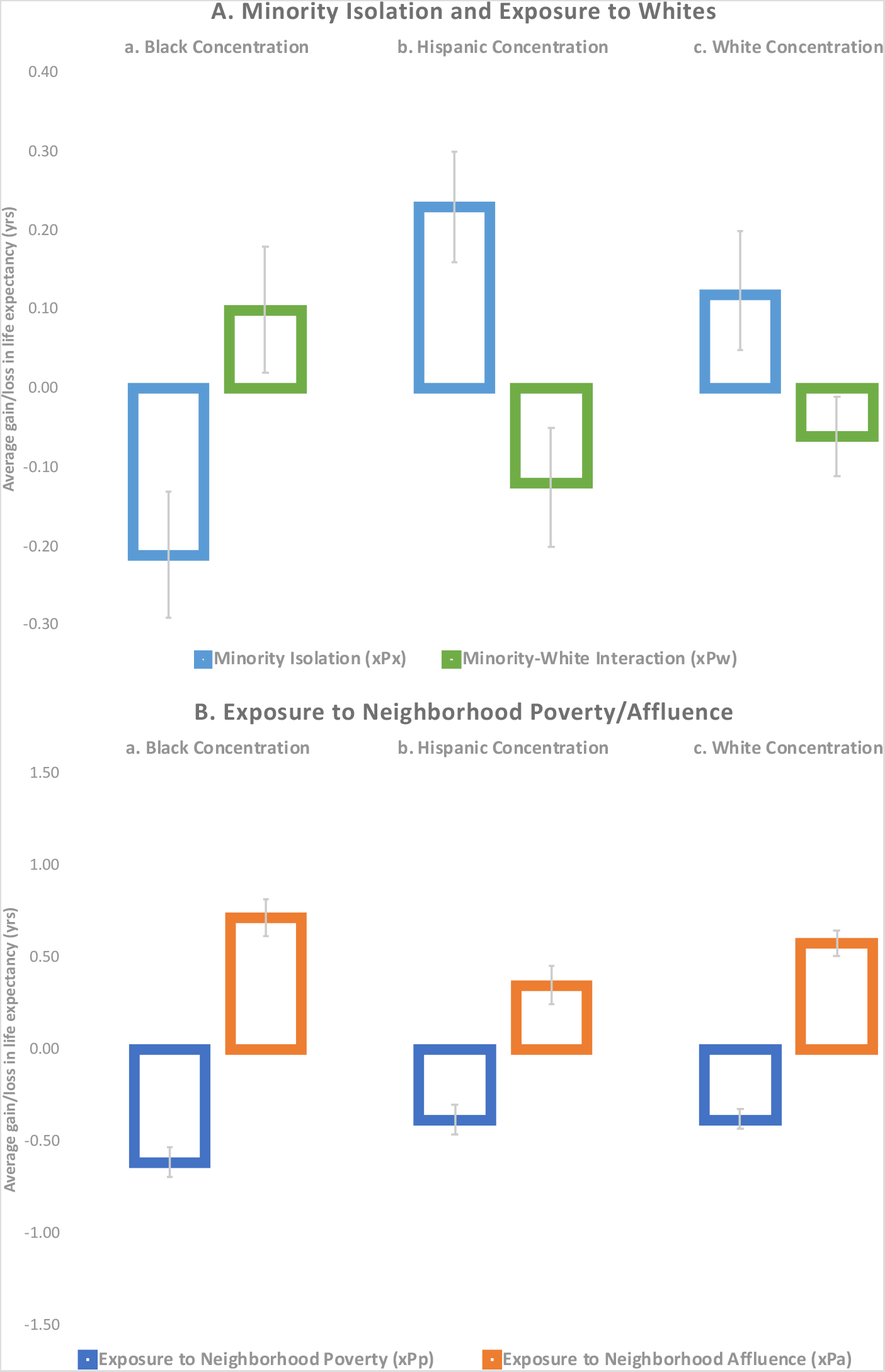
Sensitivity analysis. Contextual associations between tract probability of survival between ages 45-74 and county residential segregation, measured by minority isolation and interaction with Whites (panel A), and economic segregation measured by the interaction of race/ethnic residents with neighborhood poverty/affluence (panel B), in stratified models by race/ethnic concentration, and after accounting for county level risk factors.

**Note:** Estimates were obtained using the model specification of equation 2, where we accounted for county-level risk factors (prevalence of smoking, physical inactivity, and excessive drinking), as well as for age groups (45-54, 55-64, and 65-74) and rural/urban tract. Point estimates for all exposures and risk factors are in **Table S2**.

